# Multidimensional Sleep Health and Cognitive Risk in Midlife Primary Care: Comparing Questionnaires

**DOI:** 10.64898/2026.04.15.26350952

**Authors:** Minjee Kim, Morgan Bonham, Fangyu Yeh, Lauren Rogers, Emily H. Ho, Laura Curtis, Julia Y. Benavente, Stacy C. Bailey, Jeffrey A. Linder, Michael S. Wolf, Phyllis C. Zee

**Author notes:** **Corresponding Author:** Minjee Kim, MD, 625 N Michigan Avenue, Suite 1150, Chicago, IL, 60611.

## Abstract

**Importance:** Sleep-wake disturbances in midlife are common and potentially modifiable contributors to long-term brain health, yet primary care lacks a brief, validated tool that reliably identifies adults with early cognitive vulnerability.

**Objective:** To evaluate associations between commonly used sleep questionnaires and cognitive impairment among midlife primary care patients.

**Design, Setting, and Participants:** Cross-sectional analysis of baseline data from the MidCog cohort, an observational study of English-speaking adults aged 35 to 64 years receiving primary care at academic practices or federally qualified health centers in the Chicagoland area.

**Exposures:** Five validated sleep questionnaires were used to assess distinct sleep-wake disturbance phenotypes: (A) unsatisfactory sleep (PROMIS Sleep Disturbance T-score >55), (B) short sleep duration (<6 hours; Munich Chronotype Questionnaire), (C) obstructive sleep apnea (OSA) risk (STOP-Bang ≥3), (D) insomnia symptoms (Insomnia Severity Index ≥15), and (E) poor multidimensional sleep health (RU-SATED ≤6).

**Main Outcomes and Measures:** The primary outcome was cognitive impairment defined as an age- and education-adjusted NIH Toolbox Cognition Battery (NIHTB-CB) Fluid Composite T-score <40 (>1 SD below the population mean). Cognitive impairment defined by the Montreal Cognitive Assessment (MoCA) score <23 served as the secondary outcome. Logistic regression estimated adjusted odds ratios (aOR), controlling for age, sex, education, body mass index, hypertension, hypercholesterolemia, diabetes, smoking, depressive symptoms, and recruitment site.

**Results:** Among 646 participants (mean [SD] age, 52.3 [8.1] years; 62.4% female; 38.0% non-Hispanic Black, 38.4% non-Hispanic White, 16.0% Hispanic), cognitive impairment was present in 18.7% by NIHTB-CB and 22.3% by MoCA. Among five sleep-wake disturbance phenotypes evaluated, only poor multidimensional sleep health was consistently associated with cognitive impairment after multivariable adjustment (NIHTB-CB: adjusted OR [95% CI] = 2.03 [1.25-3.26]; MoCA: 1.98 [1.20-3.26]).

**Conclusions and Relevance:** Poor multidimensional sleep health was associated with cognitive impairment in midlife primary care patients. Brief multidimensional sleep health screening may identify individuals with early cognitive vulnerability and represent a potential strategy for targeting sleep-focused interventions to promote long-term brain health.

**Key Points:** *Question:* Among commonly used brief sleep questionnaires, which measure, if any, best identifies midlife primary care patients at risk of early cognitive vulnerability?

*Findings:* In this cross-sectional study of 646 primary care patients aged 35-64 years, poor multidimensional sleep health assessed using the RU-SATED questionnaire was the only sleep-wake disturbance phenotype consistently associated with cognitive impairment across two cognitive measures (NIH Toolbox Cognitive Battery and Montreal Cognitive Assessment).

*Meaning:* Brief multidimensional sleep health screening may help identify midlife adults with sleep-related early cognitive vulnerability in primary care and may represent a potential target for sleep-focused interventions to promote long-term brain health.

## Introduction

Sleep-wake disturbances are highly prevalent in midlife^1,2^ and associated with adverse health outcomes, including accelerated cognitive aging.^3^ Subtle cognitive changes, including declines in executive function and processing speed, often emerge during midlife and may precede clinical diagnoses by a decade or more.^4,5^ Midlife therefore represents a critical window for identifying modifiable contributors to cognitive vulnerability. However, sleep-wake disturbances frequently go underrecognized in routine primary care.

More than 150 validated sleep questionnaires exist to assess treatable sleep-wake disturbance phenotypes, including common sleep disorders (e.g., obstructive sleep apnea and insomnia) as well as suboptimal sleep characteristics (e.g., short duration or poor quality).^6^ More recently, multidimensional sleep health scales have been developed to capture multiple aspects of sleep, including regularity, satisfaction, alertness, timing, efficiency, and duration.^7^ However, it remains unclear which sleep-wake disturbance phenotypes, if any, are most informative of identifying cognitive vulnerability in midlife primary care patients. This information is necessary to prioritize targets for sleep-focused interventions.

To address this gap, we examined associations between five distinct sleep-wake disturbance phenotypes – each assessed using a brief, validated questionnaire – and cognitive impairment in a diverse cohort of midlife primary care patients.

## Methods

### Study Design and Participants

The MidCog Study is an ongoing cohort study of English-speaking adults aged 35-64 years who were recruited from 13 primary care practices across the greater Chicagoland area, including academic practices and federally qualified health centers. Individuals with severe and uncorrectable vision, hearing, or cognitive impairment (screened using the six-item screener)^8^ were excluded. Trained research staff conducted two interviews (one in-person and one telephone-based). Participants were also instructed to complete additional sleep questionnaires at home and return them to the study team using prepaid shipping materials. Details of the study protocol have been previously published.^9^ The Northwestern University Feinberg School of Medicine Institutional Review Board (STU00214736, STU00221343) approved the study, and all participants provided written informed consent.

### Questionnaire-Assessed Sleep-wake Disturbance Phenotypes

Five validated questionnaires were used to assess distinct sleep-wake disturbance phenotypes (‘sleep phenotypes’) (**Table 1**):

1. <u>Unsatisfactory sleep</u> was assessed using the Patient-Reported Outcomes Measurement Information System (PROMIS) Short Form v1.0 Sleep Disturbance 8a, an 8-item measure evaluating sleep quality and disturbances over the preceding week.^10^ Raw scores were transformed to T-scores (mean, 50; SD, 10) using the published scoring table. Unsatisfactory sleep was defined by a T-score >55, consistent with developer recommendations.
2. <u>Self-reported sleep duration</u> was assessed using the ultra-short Munich Chronotype Questionnaire (uMCTQ).^11^ Sleep duration was calculated separately for workdays and work-free days using reported sleep onset and offset times. Implausible values (sleep onset between 8:00 AM and 4:00 PM or sleep offset between 3:00 PM and 12:00 AM among non-shift workers, or sleep duration <3 or >18 hours) were individually reviewed for potential AM/PM entry errors and adjudicated by the study team.^12^ A weighted weekly average sleep duration was calculated as:

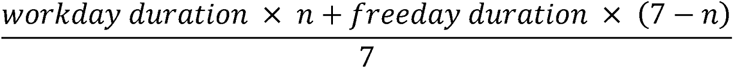 where n denotes the number of workday per week. When the number of workdays was missing, a standard 5-workday/2-freeday schedule was assumed. Short sleep duration was defined as the weighted weekly average sleep duration < 6 hours.
3. <u>Risk of obstructive sleep apnea (OSA)</u> was defined as STOP-Bang score ≥3. The STOP-Bang questionnaire includes eight dichotomous items assessing **S**noring, **T**iredness, **O**bserved apnea, high blood **P**ressure, **B**ody mass index (BMI), **a**ge, **n**eck circumference, and male **g**ender. Scores range from 0 to 8, with ≥3 indicating elevated risk of moderate-to-severe OSA and severe OSA.^13^
4. <u>Insomnia symptoms</u> were assessed using the Insomnia Severity Index (ISI), 7-item questionnaire evaluating insomnia symptoms over the preceding two weeks.^14^ Each item is scored on a 5-point Likert scale, yielding total scores from 0 to 28, with higher scores indicating more severe symptoms. Insomnia symptoms were defined using the established clinical cutoff of ≥15.^15^
5. <u>Multidimensional sleep health</u> was assessed using the RU-SATED scale.^7,16^ This 6-item questionnaire evaluates sleep **R**egularity, **S**atisfaction, **A**lertness, **T**iming, **E**fficiency, and **D**uration. Each item is rated on 3-point scale from 0 (rarely/never) to 2 (usually/always), yielding a total score ranging from 0 to 12, with higher scores indicating better sleep health.

PROMIS Sleep Disturbance and STOP-Bang were administered during the in-person and telephone interviews. The uMCTQ, ISI, and RU-SATED questionnaires were added after the initial study enrollment and were therefore available for a subset of participants. As shown in the participant flow diagram (**Figure 1**), the present analysis included participants who completed all five sleep questionnaires.

**Table 1.** Sleep Questionnaires and Operational Definitions of Sleep-Wake Disturbance Phenotypes.

Phenotypes
| Questionnaire | Sleep phenotype | Description | Threshold |
| --- | --- | --- | --- |
| PROMIS Sleep Disturbance | Unsatisfactory sleep | Assesses perceived sleep quality, depth, and restoration (8 items).<br>Scores standardized to T-scores (mean = 50, SD = 10); higher scores indicate worse sleep disturbance. | > 55 |
| Munich ChronoType Questionnaire (uMCTQ) | Short sleep duration | Assesses habitual sleep timing and duration on workdays and free days.<br>Weekly sleep duration calculated as a weighted average of workday and free-day sleep. | < 6 hours |
| STOP-Bang | Obstructive sleep apnea risk | Assesses risk of obstructive sleep apnea using 8 yes/no items.<br>Total score ranges from 0 to 8; higher scores indicate greater risk. | ≥ 3 |
| Insomnia Severity Index (ISI) | Insomnia symptoms | Assesses severity of insomnia symptoms over the past 2 weeks (7 items).<br>Total score ranges from 0 to 28; higher scores indicate more severe insomnia. | ≥ 15 |
| RU-SATED | Poor multidimensional sleep health | Assesses multidimensional sleep health across regularity, satisfaction, alertness, timing, efficiency, and duration (6 items).<br>Total score ranges from 0 to 12; lower scores indicate poorer sleep health. | ≤ 6 (lowest quartile) |

**Figure 1.**
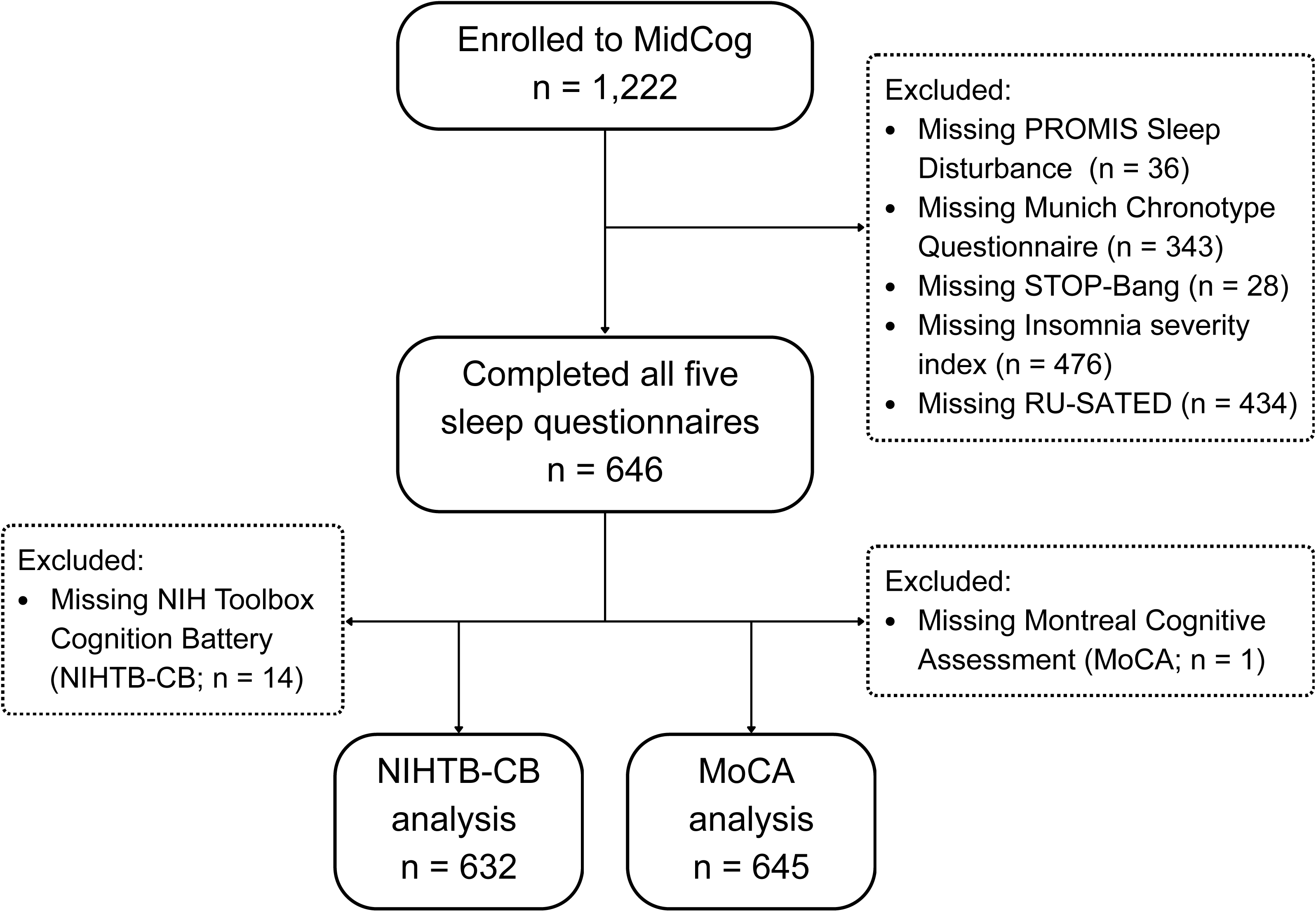
Flow Chart for Inclusion and Exclusion. The MidCog Study enrolled English-speaking adults aged 35-64 who were registered for an upcoming clinic visit within the next 6 months or has had a clinic visit in the last 18 month at one of 13 participating primary care practices across the Chicagoland area and free of vision, hearing, or cognitive impairment. Participants who completed all five sleep questionnaires were included in this study. Exclusion criteria are not mutually exclusive; participants may have been missing more than one sleep questionnaire.

### Assessment of Cognitive Impairment

Trained research staff assessed cognitive performance using two validated instruments:

#### The NIH Toolbox Cognition Battery (NIHTB-CB)

The NIHTB-CB (the primary outcome) is an iPad-based battery comprising six tests of fluid cognition (attention, executive function, processing speed, working memory, episodic memory, and associative memory) and two tests of crystallized cognition (i.e., vocabulary and reading).^17^ Because fluid cognition declines with aging and may be sensitive to early neurodegenerative processes, analyses focused on the age- and education-adjusted fluid composite score.^18^ Cognitive impairment was defined as a T-score <40 (>1 SD below the population mean). Of 646 participants, 632 completed the full NIHTB-CB battery; 14 participants were excluded from the primary outcome analysis due to incomplete administration (n = 13) and missing age-and education-adjusted T-score (n = 1). (**Figure 1**)

#### The Montreal Cognitive Assessment (MoCA)

The MoCA is a 30-point screening test of global cognition.^19^ Cognitive impairment was defined as a total score < 23.^20^ (secondary outcome) MoCA score was missing in one participant.

### Sociodemographic and Health Variables

Sociodemographic variables included age, sex, self-identified race and ethnicity (Black or African American, American Indian or Alaska Native, Hawaiian or other Pacific Islander, White or Caucasian, Asian, Hispanic/Latino/Spanish, Other/Unknown, Multiple race; collapsed into four mutually exclusive categories), and educational attainment. Health variables included body mass index (BMI; calculated as weight in kilograms divided by height in meters squared), current tobacco use (≥ 1 cigarette per day for the past 30 days), hypertension (self-reported diagnosis, systolic blood pressure ≥130 mmHg, diastolic blood pressure ≥80 mmHg, or antihypertensive medication use), hypercholesterolemia (self-reported diagnosis or cholesterol-lowering medication use), and diabetes (defined as self-reported diagnosis or antihyperglycemic medication use), and depressive symptoms (PROMIS Depression T-score >55).^22^ Recruitment sites were categorized as either academic practice or FQHC.

### Statistical Analysis

Descriptive statistics summarized baseline characteristics of the study population and by presence of each sleep phenotype. Among participants with poor multidimensional sleep health, we examined the frequency of impairment in individual RU-SATED domains (defined as item score = 0) to identify the most frequently impaired sleep dimensions.

Logistic regression was used to estimate odds ratios (ORs) for cognitive impairment associated with each sleep phenotype. Multivariable models adjusted for prespecified covariates including age, sex, education, BMI, hypertension, diabetes, hypercholesterolemia, current tobacco use, depressive symptoms, and recruitment site. For associations reaching statistical significance, we estimated age-specific marginal probabilities of cognitive impairment comparing participants with and without the sleep phenotype and evaluated potential effect modification by age, sex, recruitment site, and depressive symptoms by including interaction terms in multivariable models.

In the absence of standardized cutoff, poor multidimensional sleep health was defined as an RU-SATED total score ≤6 (range, 0-12), corresponding to the lowest quartile of the sample distribution. Sensitivity analyses modeled RU-SATED as both a continuous variable and across alternative binary thresholds to assess the stability of associations with cognitive impairment.

The primary analytic sample included participants who completed all five sleep questionnaires. To evaluate potential selection bias due to differential questionnaire completion, demographic characteristics and cognitive outcomes were compared between participants included in the analytic sample and those excluded because of missing sleep questionnaire data. Additionally, primary analyses were repeated in participants with data on at least one sleep measure (n=1,201).

Statistical significance was defined as a 2-sided p-value <0.05. All analyses were conducted using R statistical software, version 4.5.2.^21^

## Results

The analytic sample included 646 participants (mean [SD] age, 52.3 [8.1] years; 62.4% female; 38.0% non-Hispanic Black, 38.4% non-Hispanic White, 16.0% Hispanic). Cognitive impairment was present in 18.7% (118/632) based on NIHTB-CB fluid composite score (primary) and 22.3% (144/645) based on the MoCA (secondary).

Sleep-wake disturbances were common in this cohort. Unsatisfactory sleep was present in 19.0%, short sleep duration in 6.4%, elevated risk of obstructive sleep apnea in 54.8%, insomnia symptoms in 17.3%, and poor multidimensional sleep health in 24.9% of participants. As summarized in **Table 2**, non-Hispanic Black race, lower educational attainment, diabetes, depressive symptoms, and receiving care at federally qualified health centers were associated with a higher likelihood of sleep-wake disturbances across sleep phenotypes.

**Table 2.** Participant Characteristics, Overall and by Each Sleep-Wake Disturbance Phenotype.

| Characteristic <sup>a</sup> | Overall<br>N = 646 | Unsatisfactory<br>sleep <sup>b</sup><br>n = 123 (19%) | Short sleep<br>(< 6 hours) <sup>c</sup><br>n = 41 (6%) | Sleep apnea<br>risk <sup>d</sup><br>n = 354 (55%) | Insomnia<br>symptoms <sup>e</sup><br>n = 112 (17%) | Poor sleep<br>health <sup>f</sup><br>n = 161 (25%) |
| --- | --- | --- | --- | --- | --- | --- |
| Age, years | 52.3 (8.1) | 52.3 (8.5) | 53.3 (8.3) | 53.8 (7.6) § | 51.4 (8.4) | 51.9 (8.2) |
| Female | 403 (62.4%) | 88 (71.5%) † | 26 (63.4%) | 164 (46.3%) § | 71 (63.4%) | 103 (64.0%) |
| Race and ethnicity |  | † | § | § | § | § |
| *Non-Hispanic White* | 248 (38.4%) | 37 (30.1%) | 4 (9.8%) | 132 (37.3%) | 27 (24.1%) | 29 (18.0%) |
| *Non-Hispanic Black* | 245 (38.0%) | 59 (48.0%) | 30 (73.2%) | 157 (44.4%) | 52 (46.4%) | 96 (59.6%) |
| *Hispanic* | 103 (16.0%) | 23 (18.7%) | 7 (17.1%) | 47 (13.3%) | 27 (24.1%) | 29 (18.0%) |
| *Non-Hispanic Other* | 49 (7.6%) | 4 (3.3%) | 0 (0.0%) | 18 (5.1%) | 6 (5.4%) | 7 (4.3%) |
| Body mass index,<br>kg/m<sup>2</sup> | 30.5 (8.0) | 31.1 (8.2) | 34.8 (9.7) ‡ | 33.1 (8.8) § | 32.4 (8.9) † | 33.1 (9.1) § |
| Education |  | § | § | § | § | § |
| *High school or less* | 118 (18.3%) | 36 (29.3%) | 7 (17.1%) | 78 (22.0%) | 31 (27.7%) | 49 (30.4%) |
| *Some college* | 142 (22.0%) | 34 (27.6%) | 19 (46.3%) | 93 (26.3%) | 36 (32.1%) | 59 (36.6%) |
| *College graduate* | 386 (59.8%) | 53 (43.1%) | 15 (36.6%) | 183 (51.7%) | 45 (40.2%) | 53 (32.9%) |
| Hypertension | 363 (56.2%) | 81 (65.9%) † | 29 (70.7%) | 258 (72.9%) § | 76 (67.9%) ‡ | 113 (70.2%) § |
| Diabetes | 122 (18.9%) | 33 (26.8%) † | 18 (43.9%) § | 96 (27.1%) § | 31 (27.7%) ‡ | 50 (31.1%) § |
| Hypercholesterolemia | 240 (37.2%) | 49 (39.8%) | 20 (48.8%) | 177 (50.0%) § | 48 (42.9%) | 68 (42.2%) |
| Current tobacco use | 75 (11.6%) | 25 (20.3%) § | 8 (19.5%) | 52 (14.7%) ‡ | 27 (24.1%) § | 37 (23.0%) § |
| Depressive<br>symptoms<sup>g</sup> | 171 (26.5%) | 60 (49.2%) § | 16 (40.0%) † | 113 (32.0%) § | 62 (55.9%) § | 67 (41.9%) § |
| Recruitment site<sup>h</sup> |  | ‡ | § | † | § | § |
| *Academic* | 477 (73.8%) | 77 (62.6%) | 21 (51.2%) | 248 (70.1%) | 66 (58.9%) | 86 (53.4%) |
| *FQHC* | 169 (26.2%) | 46 (37.4%) | 20 (48.8%) | 106 (29.9%) | 46 (41.1%) | 75 (46.6%) |

<sup>a</sup> Mean (SD); n (%); † p<0.05, ‡ p<0.01, § p<0.001

| **Characteristic<sup>a</sup>** | **Overall**<br>**N = 646** | **Unsatisfactory**<br>**sleep<sup>b</sup>**<br>**n = 123 (19%)** | **Short sleep**<br>**(< 6 hours)<sup>c</sup>**<br>**n = 41 (6%)** | **Sleep apnea**<br>**risk<sup>d</sup>**<br>**n = 354 (55%)** | **Insomnia**<br>**symptoms<sup>e</sup>**<br>**n = 112 (17%)** | **Poor sleep**<br>**health<sup>f</sup>**<br>**n = 161 (25%)** |
| --- | --- | --- | --- | --- | --- | --- |

<sup>b</sup> Defined as Patient-Reported Outcomes Measurement Information System (PROMIS) Sleep Disturbance T-score > 55.
<sup>c</sup> Weighted weekly average sleep duration was calculated as [workday sleep duration x n + freeday sleep duration x (7 - n)] / 7 where n = number of workdays per week (5 if missing), using the ultra-short Munich ChronoType Questionnaire (uMCTQ).
<sup>d</sup> Defined as STOP-Bang score ≥ 3.
<sup>e</sup> Defined as Insomnia Severity Index (ISI) score ≥ 15.
<sup>f</sup> Defined as RU-SATED total score in the lowest quartile (≤ 6).
<sup>g</sup> Defined as PROMIS Depression T-score > 55.
<sup>h</sup> Participants were recruited from where they receive primary care (Academic Internal Medicine practices or Federally Qualified Health Centers (FQHCs)).

The median self-reported sleep duration was 7.8 hours (IQR: 7.0–8.5). Among participants with poor multidimensional sleep health (n = 161), the most frequently impaired dimension was satisfaction (52.2%), followed by efficiency (42.2%), duration (37.9%), regularity (31.7%), alertness (28.0%), and timing (19.9%).

### Associations between Sleep-Wake Disturbance Phenotypes and Cognitive Impairment

In unadjusted analyses, insomnia symptoms (NIHTB-CB: OR, 1.88 [95% CI, 1.15–3.01]; MoCA: 2.07 [1.32–3.21]) and poor multidimensional sleep health (NIHTB-CB: OR, 3.11 [2.03–4.74]; MoCA: 3.77 [2.53–5.61]) were associated with cognitive impairment. Short sleep was associated with cognitive impairment by NIHTB-CB only (OR, 2.15 [1.05–4.21]), whereas unsatisfactory sleep (OR, 2.35 [1.52–3.60]) and obstructive sleep apnea risk (OR, 1.50 [1.03–2.20]) were associated with only MoCA (**Figure 2**).

**Figure 2.**
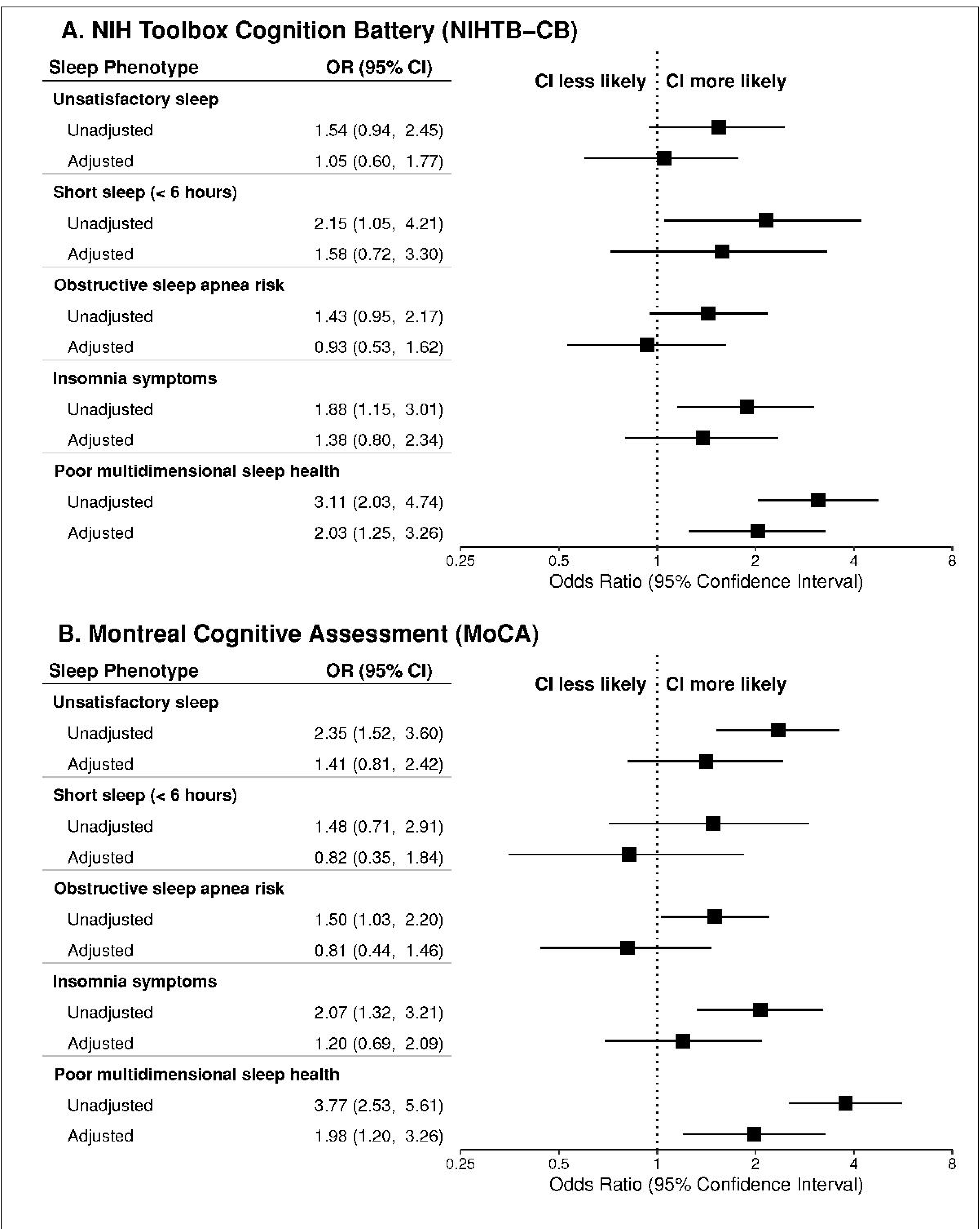
Associations between Sleep-Wake Disturbance Phenotypes and Cognitive Impairment. Associations between sleep-wake disturbance phenotypes and cognitive impairment, defined by (A) NIH Toolbox Cognition Battery (NIHTB-CB) fluid composite age-and education-adjusted T-score < 40 (primary outcome; i.e., at least one standard deviation below the population mean) or (B) the Montreal Cognitive Assessment (MoCA) < 23 (secondary outcome). Unsatisfactory sleep was defined as PROMIS Sleep Disturbance T-score > 55. Sleep duration was calculated as ([workday sleep duration x n + freeday x (7 – n)] / 7) where n = number of workdays per week (if missing, n = 5), using the ultra-short Munich ChronoType Questionnaire. Obstructive sleep apnea risk was defined as STOP-Bang score ≥ 3. Insomnia symptoms were defined as Insomnia Severity Index ≥ 15. Poor multidimensional sleep health was defined as RU-SATED total score in the lowest quartile (≤ 6). Each model included a single sleep phenotype and adjusted for age, sex, education, body mass index, current tobacco use, hypertension, hypercholesterolemia, diabetes, depressive symptoms (defined as PROMIS Depression T-score > 55), and recruitment site.

In multivariable models adjusting for sociodemographic and health characteristics, poor multidimensional sleep health was the only sleep phenotype consistently associated with cognitive impairment across both cognitive measures (NIHTB-CB: adjusted OR, 2.03 [95% CI, 1.25–3.26]; MoCA: 1.98 [1.20–3.26]) (**Figure 2**). Other sleep phenotypes, including unsatisfactory sleep, sleep duration, and insomnia symptoms, were not significantly associated with cognitive impairment. No evidence of effect modification was observed by age, sex, recruitment site, or depressive symptoms (interaction terms not significant).

To contextualize the magnitude of these associations, we estimated marginal predicted probabilities of cognitive impairment across age, comparing participants with normal versus poor multidimensional sleep health (**Supplementary Figure S1**). At age 50 years, the predicted probability of cognitive impairment was 11.2% [95% CI, 7.1–17.4%] among participants with normal sleep health and 20.8% [12.4–32.7%] among those with poor sleep health.

### Robustness Checks

When the RU-SATED total score was treated as a continuous variable, lower scores (worse sleep health) were associated with higher odds of cognitive impairment (NIHTB-CB: aOR = 1.24 [1.12–1.37], p<0.001; MoCA: aOR = 1.18 [1.07–1.31], p=0.001). Across alternative binary thresholds (RU-SATED ≤4 through ≤8), associations with the primary outcome (NIHTB-CB fluid composite T score <40) were statistically significant at thresholds ≤5 through ≤8 (aORs, 1.43–2.71), demonstrating a consistent pattern across thresholds (**Supplementary Table S1**). Similar patterns were observed for the secondary outcome (MoCA <23), with significant associations at thresholds ≤5 through ≤7 but not at ≤8.

Participants missing Insomnia Severity Index data (n=476) did not differ from the analytic sample on any key demographic characteristic. Participants missing RU-SATED (n=434) or sleep duration (n=343) data differed in racial and ethnic composition, with higher proportions of non-Hispanic Black participants compared to the analytic sample; those missing sleep duration data also had higher mean BMI and greater prevalence of the secondary outcome (MoCA <23) (**Supplementary Table S2)**. Nonetheless, findings from sensitivity analyses including all participants with data on at least one sleep questionnaire (n=1,201) were consistent with the primary complete-case results (**Supplementary Figure S2**)

## Discussion

In this cross-sectional study of midlife primary care patients, poor multidimensional sleep health, assessed with the RU-SATED questionnaire, was the only phenotype among five commonly used brief sleep screening instruments consistently associated with cognitive impairment across two cognitive measures. These findings suggest that multidimensional sleep health screening may better capture sleep-related cognitive vulnerability in midlife than single-domain measures, and support its potential role as a practical tool for identifying patients who might otherwise go unrecognized in routine primary care. No participant carried a clinical diagnosis of cognitive impairment, consistent with evidence that subtle cognitive changes emerge in midlife and may precede clinical recognition by a decade or more.^4,5^

The independent association between multidimensional sleep health and cognitive performance, beyond that of insomnia symptoms, sleep satisfaction, or duration alone, reinforces the conceptual framework proposed by Buysse, in which sleep health reflects a composite pattern of regularity, satisfaction, alertness, timing, efficiency, and duration that promotes physical and mental well-being.^7^ Composite assessments, such as RU-SATED, have been more strongly associated with health outcomes than any single dimension, as demonstrated in the Midlife in the United States (MIDUS) study^22–24^ and others.^25,26^ Our findings extend this evidence to cognitive outcomes in a primary care setting.

The most commonly impaired RU-SATED dimensions in our sample were satisfaction, efficiency, duration, and regularity. This pattern mirrors findings from Smith et al., who identified short, dissatisfied, inefficient, and irregular sleep as the predominant unhealthy sleep phenotype, affecting 27% of working U.S. adults in midlife.^27^ Such consistency underscores the value of assessing sleep health as a multidimensional construct and suggests potential intervention targets. Recent trial evidence supports this direction: individualized sleep counseling in habitual short sleepers improved not only sleep duration but also improved satisfaction and daytime alertness.^28^

Our findings align with prior research linking sleep-wake disturbances to cognitive decline in older adults,^29^ although relatively few studies have examined these relationships in midlife. Nearly one in five participants in our cohort demonstrated fluid cognition scores more than one standard deviation below age- and education-adjusted norms – a marker of cognitive vulnerability relevant to dementia risk. The mechanisms underlying the observed association between sleep health and cognitive function are likely multifactorial and bidirectional and may involve impaired glymphatic clearance of metabolic waste,^30,31^ altered brain glucose metabolism,^32^ systemic inflammation,^33^ and imbalanced autonomic nervous system.^34^ Conversely, early cognitive impairment may itself disrupt circadian rhythms and sleep architecture, suggesting a reinforcing cycle that warrants longitudinal investigation.

Clinically, brief tools such as RU-SATED may help primary care clinicians identify patients with early cognitive vulnerability who might otherwise go unrecognized. Unlike objective sleep assessments such as actigraphy or polysomnography, which require specialized equipment, staffing, and data-processing infrastructure, RU-SATED can be self-administered or completed during rooming in approximately one minute, scored immediately, and integrated into existing electronic health record workflows.^35^ A positive screen could prompt further cognitive evaluation or referral to sleep medicine, providing a low-burden entry point for risk stratification. Although standardized clinical thresholds for RU-SATED have not yet been established, the consistent associations observed across a range of binary thresholds suggest that even conservative cutoffs may be informative in primary care setting.

Notably, short sleep duration (<6 hours) was not associated with cognitive impairment in our sample. This is consistent with some prior studies^23,36^ but not others,^37,38^ and may reflect differences in measurement approach, thresholds, or sample characteristics. The mixed literature^39^ further suggests that sleep duration alone may not adequately capture the complexity of sleep-wake patterns relevant to cognitive aging.

Poor sleep health was more prevalent among participants receiving care at federally qualified health centers (FQHCs) and among those identifying as non-Hispanic Black. FQHCs predominantly serve low-income and historically minoritized populations – groups that bear a disproportionate burden of both poor sleep health and dementia risk, yet face the greatest barriers to sleep specialty care.^40–42^ These disparities likely reflect the cumulative effects of structural racism, neighborhood-level environmental stressors (e.g., noise, light pollution, perceived safety), economic precarity, and chronic psychosocial stress on sleep-wake regulation.^43^ Cognitive vulnerability in these populations is further compounded by limited access to neuropsychological evaluation and specialty referral. Any future intervention research should prioritize recruitment from these populations and evaluate equity outcomes explicitly; RU-SATED’s brevity and cross-cultural validation make it a reasonable candidate for testing in diverse primary care settings, but its utility as an equity-relevant screening tool requires prospective evaluation.

This study has several limitations. First, the cross-sectional design precludes causal inference. Second, sleep phenotypes were captured using questionnaires rather than objective measures such as actigraphy or polysomnography, introducing potential recall or misclassification bias; however, questionnaires remain the most practical approach for routine primary care screening. Third, the requirement for complete data across all five sleep questionnaires resulted in exclusion of approximately half the enrolled sample. Participants missing the RU-SATED and sleep duration data differed from the analytic sample on race and ethnicity composition, with non-Hispanic Black participants overrepresented among non-completers. Nonetheless, sensitivity analyses including all participants with data on at least one sleep measure yielded consistent findings, suggesting the complete-case restriction did not materially bias the primary associations. Fourth, the study-defined RU-SATED threshold (≤6; lowest quartile) was derived from this sample’s distribution rather than a pre-specified or externally validated cutoff, which may influence the observed association. However, sensitivity analyses modeling RU-SATED as a continuous variable and across multiple binary thresholds yielded consistent associations with cognitive impairment, indicating that the observed relationship was not dependent on the specific cutoff used. Establishing standardized clinical cutoffs and validating screening performance across diverse clinical settings should be a priority for future research. Fifth, the exclusion of non-English speakers limits generalizability to non-English-speaking patients, including a substantial proportion of Hispanic adults in the Chicagoland primary care population.

Despite these limitations, this study provides empirical guidance on instrument selection for future sleep-targeted intervention research. Sleep-wake disturbances are highly prevalent yet underrecognized in routine care, and it remains unclear which dimensions of sleep health should be prioritized or how to best identify individuals at greatest risk. Pragmatic trials embedding multidimensional sleep health assessments into primary care workflows — leveraging electronic health record–integrated tools and team-based care — represent a promising next step. Whether interventions targeting the most commonly impaired sleep dimensions (satisfaction, efficiency, duration, regularity) translate into long-term cognitive benefit should be a priority for future research.

## Conclusion

Poor multidimensional sleep health was associated with early cognitive vulnerability in midlife primary care patients. Brief multidimensional sleep health screening may help identify individuals at risk who might otherwise go unrecognized and represents a potentially scalable strategy for targeting sleep-focused interventions. Future work should investigate whether addressing poor sleep health can prevent or delay cognitive decline, particularly in high-risk populations.

## Data Sharing Statement

The de-identified individual participant data that underlie the results reported in this article will be made available to qualified researchers upon reasonable request.

## Acknowledgments

We thank the MidCog study participants and research staff. MK and FY had full access to all the data in the study and take responsibility for the integrity of the data and the accuracy of the data analysis.

M.K. received research funding from Genentech, Inc. on behalf of the institution and received a consulting fee from Hinshaw & Culbertson LLP. All other authors report no conflict of interest.

