## Supplementary material for "Multidimensional Sleep Health and Cognitive Risk in Midlife Primary Care: Comparing Questionnaires": Figure S1, Table S1, Table S2, Figure S2

### Contents:

**Figure S1.** Age-specific Marginal Predicted Probabilities of Cognitive Impairment by Multidimensional Sleep Health Status

**Table S1.** Discriminative Performance and Associations Between RU-SATED and Cognitive Impairment Across Alternative Thresholds

**Table S2.** Baseline Characteristics of the Analytic Sample and Participants Excluded Due to Missing Sleep Questionnaire Data

**Figure S2:** Associations between Sleep Phenotypes and Cognitive Impairment among Participants with Data on at least One Sleep Questionnaire (n = 1,201)

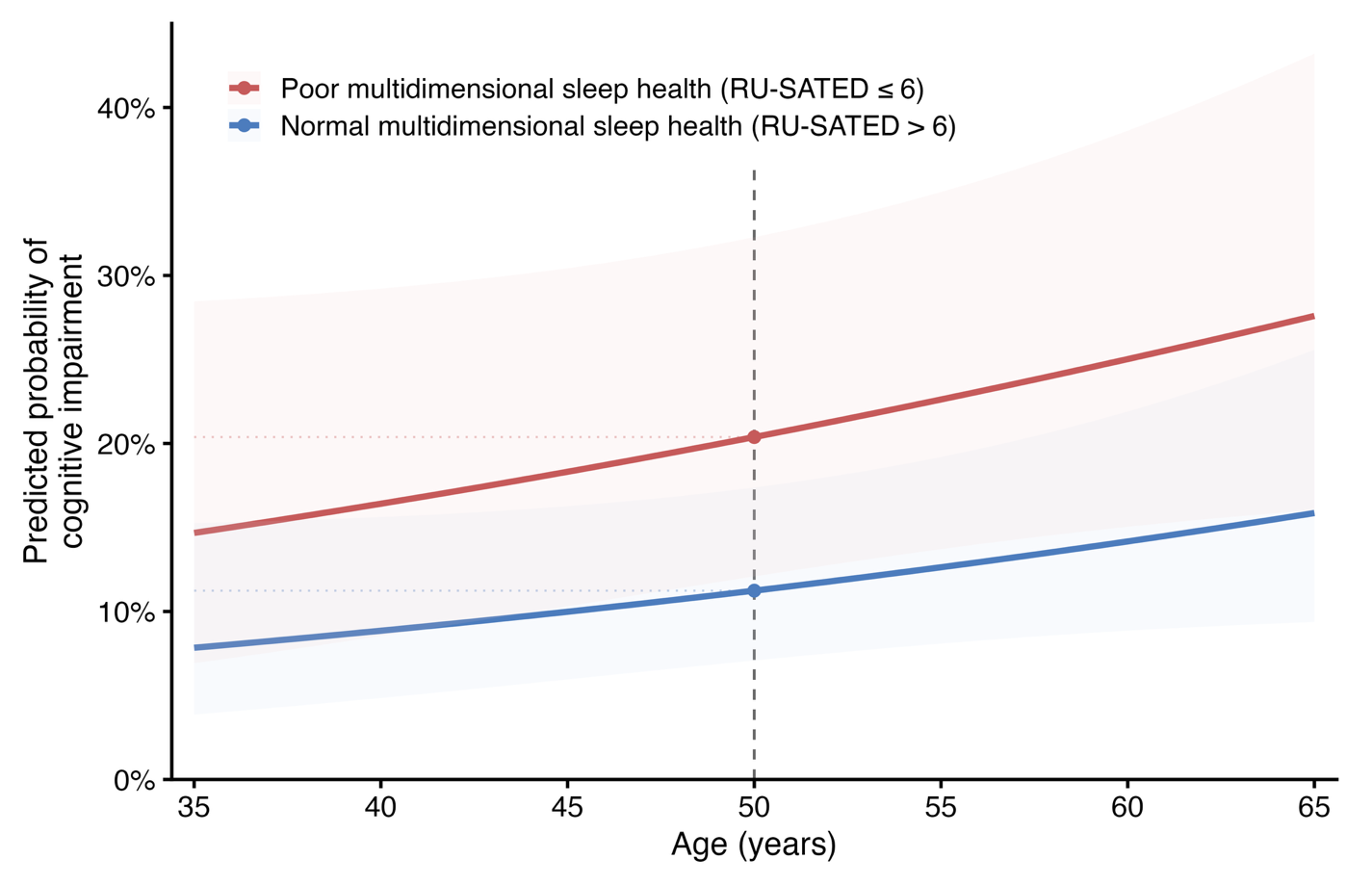
**Figure S1.** Age-specific Marginal Predicted Probabilities of Cognitive Impairment by Multidimensional Sleep Health Status

Age-specific marginal predicted probabilities of cognitive impairment (NIHTB-CB fluid composite T-score <40), comparing participants with poor multidimensional sleep health (RU-SATED ≤6) and normal sleep health (RU-SATED >6). Estimates were derived from multivariable logistic regression models adjusting for age, sex, education, body mass index, hypertension, diabetes, hypercholesterolemia, current tobacco use, depressive symptoms, and recruitment site, averaging over the observed covariate distribution. Shaded areas represent 95% confidence intervals. The vertical dashed line denotes age 50 years as an illustrative reference point.

**Table** **S1**. Discriminative Performance and Associations Between RU-SATED and Cognitive Impairment Across Alternative Thresholds

|  |  | **Discriminative Performance** | | | **Association with Cognitive Impairment, Adjusted OR (95% CI), pᵃ** | |
| --- | --- | --- | --- | --- | --- | --- |
| **RU-SATED Threshold** | **n (%)** | **AUC** | **Sensitivity** | **Specificity** | **NIHTB-CB (Primary)** | **MoCA (Secondary)** |
| Continuous (per 1-pt decrease) | — | 0.698 | — | — | 1.24 (1.12–1.37) p<0.001 | 1.18 (1.07–1.31) p=0.001 |
| ≤ 4 | 54 (8.4%) | 0.538 | 14.4% | 93.2% | 1.43 (0.71–2.78) p=0.304 | 1.57 (0.77–3.15) p=0.211 |
| ≤ 5 | 99 (15.3%) | 0.584 | 28.8% | 87.9% | 1.95 (1.14–3.28) p=0.013 | 2.10 (1.20–3.67) p=0.009 |
| **≤ 6**^b^ | **161 (24.9%)** | **0.619** | **44.1%** | **79.8%** | **2.03 (1.25–3.26) p=0.004** | **1.98 (1.20–3.26) p=0.008** |
| ≤ 7 | 235 (36.4%) | 0.633 | 57.6% | 68.9% | 2.11 (1.33–3.35) p=0.001 | 1.62 (1.00–2.63) p=0.048 |
| ≤ 8 | 339 (52.5%) | 0.649 | 76.3% | 53.5% | 2.71 (1.65–4.55) p<0.001 | 1.61 (0.96–2.69) p=0.070 |

RU-SATED scores range from 0 to 12, with lower scores indicating worse sleep health.

Abbreviations: AUC, area under the receiver operating characteristic curve; MoCA, Montreal Cognitive Assessment; NIHTB-CB, NIH Toolbox Cognition Battery; OR, odds ratio.

ᵃ Adjusted for age, sex, education, body mass index, hypertension, hypercholesterolemia, diabetes, current tobacco use, depressive symptoms (PROMIS Depression T-score >55), and recruitment site.

^b^ Bold row (≤6) indicates the study-defined threshold, corresponding to the lowest quartile of the sample distribution.

**Table S2.** Baseline Characteristics of the Analytic Sample and Participants Excluded Due to Missing Questionnaire Data

| **Characteristic** | **Analytic sample N = 646^1^** | **Missing ISI N = 476^1^** | **Missing RU-SATED N = 434^1^** | **Missing uMCTQ N = 343^1^** | **Missing PROMIS N = 36^1^** | **Missing STOP-Bang N = 28^1^** |
| --- | --- | --- | --- | --- | --- | --- |
| Age, years | 52.3 (8.1) | 52.3 (8.3) | 52.8 (8.1) | 51.8 (8.4) | 50.0 (8.7) | 50.8 (8.4) |
| Female | 403 (62.4%) | 284 (59.8%) | 257 (59.4%) | 193 (56.4%) | 18 (51.4%) | 12 (42.9%) † |
| Race and ethnicity |  |  | † | † | ‡ | ‡ |
| *Non-Hispanic White* | 248 (38%) | 179 (38%) | 161 (38%) | 111 (33%) | 8 (23%) | 5 (19%) |
| *Non-Hispanic Black* | 245 (38%) | 208 (44%) | 191 (45%) | 166 (49%) | 22 (63%) | 18 (67%) |
| *Hispanic* | 103 (16%) | 52 (11%) | 45 (11%) | 43 (13%) | 1 (2.9%) | 1 (3.7%) |
| *Non-Hispanic Other* | 49 (7.6%) | 31 (6.6%) | 31 (7.2%) | 19 (5.6%) | 4 (11%) | 3 (11%) |
| Body mass index, kg/m² | 30.5 (8.0) | 30.8 (8.2) | 31.1 (8.1) | 31.7 (8.3) † | 29.9 (8.7) | 26.4 (4.0) ‡ |
| Education |  |  |  |  |  |  |
| *High school or less* | 118 (18%) | 73 (15%) | 60 (14%) | 71 (21%) | 9 (26%) | 7 (25%) |
| *Some college* | 142 (22%) | 104 (22%) | 99 (23%) | 79 (23%) | 12 (34%) | 9 (32%) |
| *College graduate* | 386 (60%) | 298 (63%) | 274 (63%) | 192 (56%) | 14 (40%) | 12 (43%) |
| Hypertension^2^ | 363 (56.2%) | 275 (57.8%) | 255 (58.8%) | 208 (60.6%) | 23 (63.9%) | 17 (60.7%) |
| Diabetes^3^ | 122 (18.9%) | 97 (20.4%) | 97 (22.4%) | 82 (23.9%) | 8 (22.2%) | 6 (21.4%) |
| Hypercholesterolemia^4^ | 240 (37.2%) | 176 (37.0%) | 171 (39.4%) | 131 (38.2%) | 12 (33.3%) | 11 (39.3%) |
| Current tobacco use | 75 (11.6%) | 47 (10.5%) | 44 (10.9%) | 45 (14.2%) | 0 (0.0%) | 0 (0%) |
| Depressive symptoms^5^ | 171 (26.5%) | 110 (24.7%) | 101 (25.0%) | 85 (26.8%) | 1 (33.3%) | 0 (0%) |
| NIHTB-CB T-score < 40^6^ | 118 (18.7%) | 99 (21.7%) | 87 (20.8%) | 71 (21.6%) | 9 (27.3%) | 8 (30.8%) |
| MoCA < 23^7^ | 144 (22.3%) | 121 (25.9%) | 109 (25.5%) | 96 (28.5%) † | 13 (39.4%) † | 9 (33.3%) |
| Recruitment site^8^ |  |  | † |  | ‡ |  |
| *Academic* | 477 (74%) | 373 (78%) | 344 (79%) | 239 (70%) | 19 (53%) | 17 (61%) |
| *FQHC* | 169 (26%) | 103 (22%) | 90 (21%) | 104 (30%) | 17 (47%) | 11 (39%) |
| Abbreviations: ISI = Insomnia Severity Index; RU-SATED = Regularity, Satisfaction, Alertness, Timing, Efficiency, Duration; uMCTQ = ultra-short Munich Chronotype Questionnaire; PROMIS = Patient-Reported Outcomes Measurement Information System Sleep Disturbance; STOP-Bang = Snoring, Tiredness, Observed apnea, blood Pressure, Body mass index, Age, Neck circumference, Gender.  ^1^Mean (SD); n (%)  † p<0.05, ‡ p<0.01, § p<0.001, using Wilcoxon rank sum test (continuous) or Pearson's Chi-squared test (categorical); Interpret with caution for PROMIS and STOP-Bang given small excluded group sizes (n<40). | | | | | | |
| ^2^ Defined as self-reported diagnosis, systolic blood pressure ≥130 mmHg, diastolic blood pressure ≥80 mmHg, or antihypertensive medication use. | | | | | | |
| ^3^ Defined as self-reported diagnosis or antihyperglycemic medication use. | | | | | | |
| ^4^ Defined as self-reported diagnosis or cholesterol-lowering medication use. | | | | | | |
| ^5^ Defined as PROMIS Depression T-score > 55. | | | | | | |
| ^6^ NIH Toolbox Cognition Battery Fluid Composite age- and education-adjusted T-score (mean = 50, SD = 10) < 40. | | | | | | |
| ^7^ Montreal Cognitive Assessment total score (range: 0-30) < 23. | | | | | | |
| ^8^ Participants were recruited from academic internal medicine practices or Federally Qualified Health Centers (FQHC). | | | | | | |

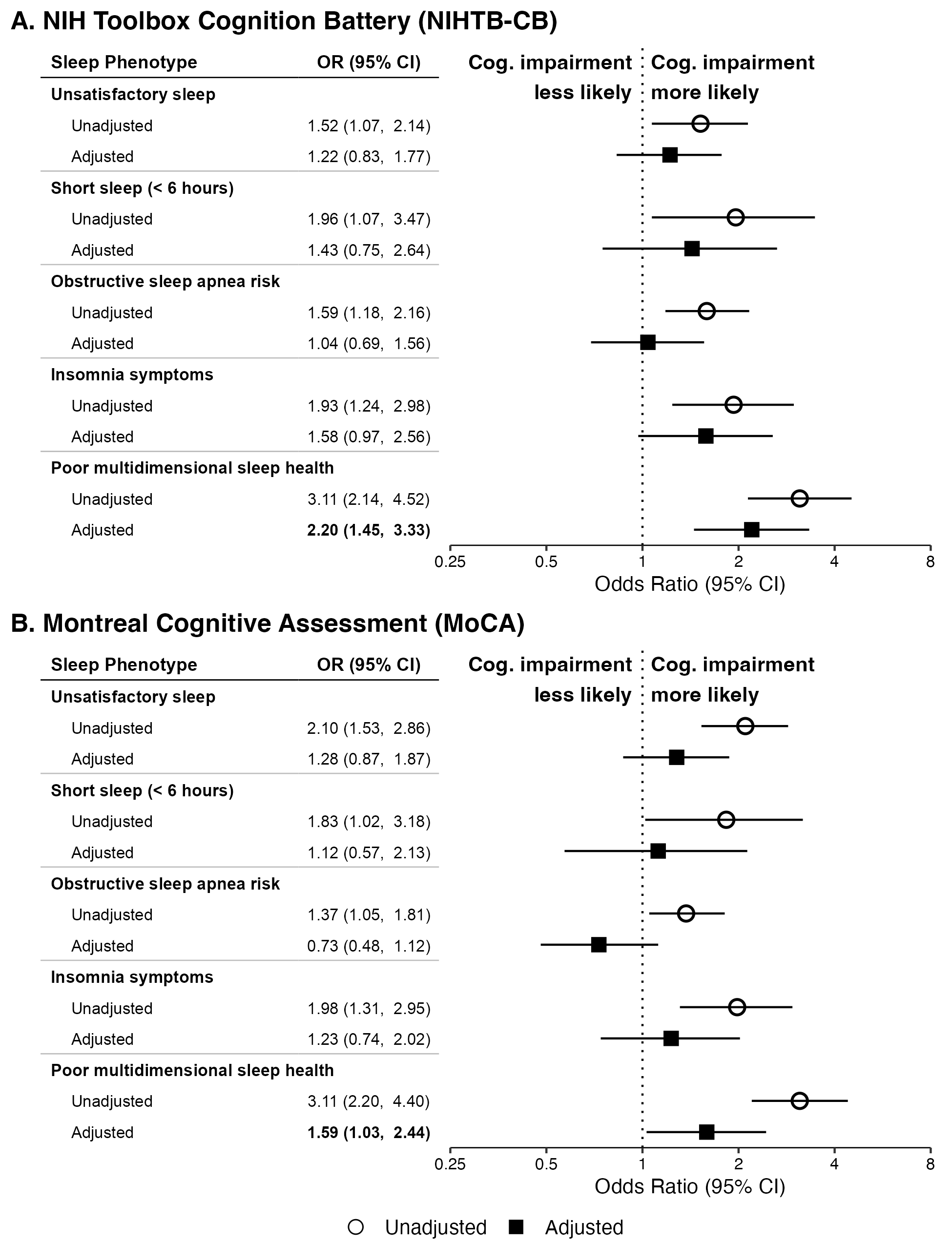
**Figure S2:** Associations between Sleep Phenotypes and Cognitive Impairment among Participants with Data on at least One Sleep Questionnaire (n = 1,201)

Forest plots show unadjusted and adjusted odds ratios (ORs) with 95% confidence intervals (CIs) for the association between each sleep phenotype and cognitive impairment, separately for the NIH Toolbox Cognition Battery fluid composite T-score <40 (Panel A) and Montreal Cognitive Assessment score <23 (Panel B). Adjusted models included age, sex, education, body mass index, hypertension, hypercholesterolemia, diabetes, current tobacco use, depressive symptoms (PROMIS Depression T-score >55), and recruitment site. Bold text indicates statistical significance (95% CI excludes 1.0). The reference line is at OR = 1.0.

Abbreviations: CI, confidence interval; ISI, Insomnia Severity Index; MoCA, Montreal Cognitive Assessment; NIHTB-CB, NIH Toolbox Cognition Battery Fluid Composite; OR, odds ratio; PROMIS, Patient-Reported Outcomes Measurement Information System; RU-SATED, Regularity, Satisfaction, Alertness, Timing, Efficiency, Duration; STOP-Bang, Snoring, Tiredness, Observed apnea, blood Pressure, Body mass index, Age, Neck circumference, Gender; uMCTQ, ultra-short Munich Chronotype Questionnaire.
